# Breastfeeding duration is associated with larger cortical gray matter volumes in children from the ABCD study

**DOI:** 10.1101/2022.05.23.22274926

**Authors:** Christian Núñez, Alfredo García-Alix, Gemma Arca, Thais Agut, Nuria Carreras, Maria J. Portella, Christian Stephan-Otto

## Abstract

Despite the numerous studies in favor of breastfeeding for its benefits in cognition and mental health, the long-term effects of breastfeeding on brain structure are still largely unknown. Here we analyzed 7,860 MRI images of children 9 to 11 years of age from the Adolescent Brain Cognitive Development (ABCD) dataset in order to study the relationship between breastfeeding duration and cerebral gray matter volumes. We also explored the potential mediatory role of brain volumes on behavior. Whole-brain voxel-based morphometry analyses revealed an association mainly between breastfeeding duration and larger bilateral volumes of the pars orbitalis and the lateral orbitofrontal cortex. In particular, the association with the left pars orbitalis and the lateral orbitofrontal cortex proved to be very robust, and it appeared to mediate the relationship between breastfeeding duration and the negative urgency dimension of the UPPS-P Impulsive Behavior Scale. Global gray matter volumes were also significant mediators for behavioral problems as measured with the Child Behavior Checklist. Overall, our findings suggest that breastfeeding is an important factor in the proper development of the brain, particularly for the pars orbitalis and lateral orbitofrontal regions. This, in turn, may impact impulsive personality and mental health in early puberty.

## Introduction

The effects of breastfeeding on cognition and intelligence have been broadly examined, and most studies report a beneficial effect of breastfeeding in this regard [see Horta et al.^1^ for a meta-analysis]. Breastfeeding also seems to come with additional general health benefits, such as reduced mortality in children and protection against obesity and other diseases.^2^ Some reports also point to a link between breastfeeding and better preserved mental health later in life,^3^ including less risk of developing schizophrenia,^4,5^ attention deficit hyperactivity disorder,^6^ or severe depressive symptoms.^7^ Despite all the evidence in favor of breastfeeding, on average only 1 in 4 infants from high-income countries are still breastfeeding at 12 months of age.^2^

In contrast to the wealth of literature available on the effects of breastfeeding on cognition and intelligence, only a limited number of studies have explored the relationship between breastfeeding and brain structure, and only a handful of magnetic resonance imaging (MRI) studies have examined the association between breastfeeding and gray matter (GM) volume. It has generally been found that breastfeeding is associated with larger GM volume in children.^8–12^ Using a dichotomous measure, breastfed participants showed larger global^8^ and localized GM volumes, specifically the left inferior temporal and superior parietal.^9^ Duration of breastfeeding has also been associated with larger volumes of the bilateral hippocampi,^12^ the inferior frontal gyrus and rolandic operculum,^10^ and the striatum and medial orbital gyrus.^11^ Other studies included only participants born preterm, such as that by Belfort et al.,^13^ who found positive associations between breastfeeding and volumes of the hippocampus and deep GM nuclei at term-equivalent age, even though these associations seemed to disappear at 7 years of age. Similarly, Ottolini et al.,^14^ also examining MRI acquired at term-equivalent age, found larger volumes in a region encompassing the hippocampus and the amygdala in breastfed infants. Other preterm studies employing more global GM measures were unable to find any positive association with breastfeeding at term-equivalent age^15^ or in adolescence.^16^ Notably, an experimental study with macaque monkeys found that breastfeeding promoted maturation of the cortex.^17^ Moreover, other studies have generally observed positive associations between breastfeeding and white matter integrity and cerebral connectivity,^14–21^ and with cortical thickness.^22^

It is important to note some limitations that are present in most or all of the aforementioned studies on breastfeeding and GM volumes. First, only three of them^9–11^ employed a whole-cortex approach, by means of voxel-based morphometry (VBM) techniques, while the rest employed global, nonspecific GM measures or else examined only specific, predefined structures (region of interest analysis), therefore not producing a complete picture of the effects of breastfeeding on the brain. Second, most of them used categorical or even dichotomous (yes/no), rather than continuous, variables to characterize breastfeeding, thus limiting the ability to obtain more accurate results. Third, the samples employed were relatively small, with the study of Koshiyama et al.^11^ being the largest with 207 participants.

Here we aimed to analyze the association between the duration of breastfeeding and cerebral GM volumes in early puberty, by means of whole-brain VBM techniques, employing the very large Adolescent Brain Cognitive Development (ABCD) dataset, which recruited and acquired MRI images from more than 11,000 children.^23^ A recent publication using data from the ABCD database showed beneficial effects of breastfeeding on cognition,^24^ which is in line with previous studies. We expected to find a positive association between duration of breastfeeding and GM volumes. However, given the paucity of studies on this topic, we did not set any hypothesis about which specific brain regions would show larger GM volumes. We also aimed to explore the possible mediatory role of GM volumes on cognition, mental health, and other behavioral measures.

## Methods

### Participants

A total of 7,860 participants from the ABCD 3.0 dataset were included in the analyses [median (IQR) age = 119 (14) months; 3,920 females). Of the initial sample of 11,230 children who had a recommended (ABCD-validated) T1-weighted brain image, we excluded 3,370 for the following reasons: 763 for concerns about the resulting images after the segmentation procedure (see the “Neuroimaging data acquisition and processing” section below); 410 that were referred to a doctor after radiological assessment or for whom radiological assessment was not possible; 366 for presenting a major medical condition (e.g., autism, epilepsy, etc.) but still considered eligible for participation in the ABCD study; 1,326 for whom pregnancy-related questions (including breastfeeding duration) were not answered by their biological mother; and 505 that had a null or missing value in the breastfeeding duration variable or in any of the other variables used in the main analysis (see the “Neuroimaging analysis” section below).

### Breastfeeding duration data

Breastfeeding duration information was obtained from the “ABCD Developmental History Questionnaire” that was completed by the mothers of the participants. The exact question that they responded to was “For how many months was your child breast fed?”. As stated before, we excluded non-biological mothers in order to maximize the accuracy of the answers. We used the continuous variable (months of breastfeeding) in the analyses. In our sample, the median (IQR) of breastfeeding duration was 6 (11) months. The minimum and maximum values of breastfeeding duration were 0 and 84 months, respectively.

### Neuroimaging data acquisition and processing

We gathered the 11,230 recommended high-resolution T1-weighted structural images acquired at the baseline visit of the participants from the ABCD 3.0 dataset. The ABCD study employed 3 different 3T scanner platforms for MRI acquisition at 22 sites throughout the United States. Information on the imaging procedures and scanning protocols may be found elsewhere.^25^ We processed and segmented all the T1-weighted images with CAT12 (v12.7, r1742; http://www.neuro.uni-jena.de/cat/), a toolbox for SPM12 (https://www.fil.ion.ucl.ac.uk/spm/software/spm12/), running under MATLAB (Release 2020b, The MathWorks, Inc., Natick, MA, USA). In order to improve the accuracy of the segmentation, we employed a custom pediatric tissue probability map that we generated from 1,000 participants’ (50% males/females) images, randomly chosen from the ABCD dataset, with the CerebroMatic Toolbox for SPM12.^26^ The segmentation procedure resulted in the generation of modulated and warped GM images for each individual, which we smoothed using a Gaussian kernel of 8 mm full width at half maximum (FWHM). We used these smoothed GM segments for the whole-brain VBM analyses. We used the Neuromorphometrics atlas (http://www.neuromorphometrics.com) as reference for our results. To ensure the quality of the data, we employed the “Data quality” tools provided in CAT12 and the reports generated after the segmentation procedure to detect participants that might have been processed inappropriately. Following this, we excluded a total of 763 participants (6.8% of the total sample).

### Neuroimaging analysis

First, we performed whole-brain VBM multiple regression analysis of the GM segments, with breastfeeding duration (in months) as the independent variable and age, sex, education level, race, ethnicity, scanner, and total intracranial volume as covariates. Second, in order to test the robustness of our result, we split our sample and repeated the previous VBM analysis in each of the two halves. To do this, we used the suggested ABCD Reproducible Matched Samples (ARMS) provided by the ABCD-BIDS Community Collection (https://collection3165.readthedocs.io/), after applying all the exclusion criteria listed previously. Third, since the ABCD dataset includes participants that are related (siblings), we repeated the original VBM analysis multiple times by randomly selecting only one sibling per family each time, to ensure that the potential genetics/shared environment effects were not confounding the results. Fourth, we repeated the original VBM analysis with the addition of several potential confounding variables, one at a time. We tested the following factors: weight at birth, prematurity, parental age at child’s birth, number of problems during pregnancy or birth, maternal tobacco or alcohol use during pregnancy, family income, parental education level, current degree of parental monitoring and attentiveness, current body mass index, and degree of pubertal development. Finally, we performed two additional variations of the original VBM analysis, one of them excluding preterm participants (born before 37 gestational weeks) and the other replacing the original scanner covariates with more precise variables representing a unique identifier for each scanner. The full list of variables of interest and covariates that we used in the VBM analyses may be found in Supplementary Table 1. In all VBM analyses, we employed a GM masking absolute threshold of 0.2, as well as a family-wise error (FWE)-corrected voxel-level threshold defined by p < 0.001 and a minimum cluster size of 200 voxels, save for the VBM analyses using split samples, in which we used a more permissive FWE-corrected voxel-level threshold defined by p < 0.05 and a minimum cluster size of 200 voxels. Overall, we employed highly restrictive thresholds to ensure that we obtained robust and scientifically meaningful results.

### Behavioral data

To address the potential mediatory role of GM volumes between breastfeeding duration and behavior, we gathered the following baseline data from the ABCD 3.0 dataset:

- Global cognition: We used the uncorrected Total Composite Score from the NIH toolbox, which assesses several cognitive domains by means of 7 tests, as a general measure of cognition.^27^
- Behavioral problems: The parent Child Behavior Checklist^28^ is composed of several items asking the parents for information about possible behavioral and emotional problems of their children. We obtained 3 raw composite scores: total problems, internalizing problems, and externalizing problems. We performed post-hoc analyses with the internalizing and externalizing scores only if a significant result with the total problems score arose.
- Prodromal psychotic experiences: The Prodromal Questionnaire–Brief Child Version^29^ asks the participants about 21 possible psychotic-like experiences. Participants indicate whether they have experienced any of these, and, if so, whether or not they were troubled by the experience and to what extent, on a 1-5 scale. We computed 2 scores: psychotic experiences (i.e., the number of psychotic-like experiences reported by the participants) and troubling psychotic experiences (i.e., the sum of all the troubling scores reported by the participants).

After reviewing our neuroimaging analyses (see “Results” section below), we decided to obtain these additional behavioral measures:

- Vocabulary: We used the uncorrected Picture Vocabulary Test Score, which is one of the tests included in the NIH toolbox, as a measure of verbal performance.^30^
- Prosociality: Three items from the prosocial scale of the Strengths and Difficulties Questionnaire^31^ were answered on a 0-2 scale by both participants and their parents. We computed the sum of the total scores from both the participants’ and the parents’ questionnaires and used this figure as a measure of prosociality.
- Impulsivity: The ABCD Version of the UPPS-P Impulsive Behavior Scale for Children-Short Form^32^ is comprised of 20 items assessing 5 different impulsivity dimensions (lack of premeditation, lack of perseverance, negative urgency, positive urgency, and sensation seeking). We computed the total impulsivity score for all 20 items as well as the total sub-scores for each dimension. We performed post-hoc analyses with the dimension sub-scores only if a significant result with the total impulsivity score arose.

Additional information about the provenance of these variables and summary measures may be found in Supplementary Table 2.

### Mediation analysis

We explored the potential mediatory role of the cerebral regions of interest, identified by means of the VBM analysis, as well as global GM volume, between breastfeeding duration and the behavioral scores mentioned in the previous paragraph. We performed the mediation analyses with the PROCESS tool for SPSS.^33^ We set a simple mediation model with one mediator and 5,000 bootstrap samples. We used breastfeeding duration as the independent variable (X), brain GM volumes as the mediator variable (M), and behavioral scores as the dependent variable (Y). For the X → M regression model we employed the same main covariates used in the neuroimaging analysis. For the M → Y and the X → Y (total effect) regression models, we replaced the scanner variables with dummy variables to represent each of the 22 sites where participants were recruited. The standardized coefficients (β) are reported. We estimated the p-values for the indirect effects with the calculator provided by Falk and Biesanz.^34^

## Results

### Whole-brain VBM analysis on breastfeeding duration

The main VBM analysis revealed breastfeeding duration to be associated with larger bilateral GM volumes at the orbital part of the inferior frontal gyrus (pars orbitalis) and the lateral orbitofrontal cortex, among other adjacent regions. We found an additional association with a smaller cluster located at the left postcentral gyrus and the superior parietal lobule (Figure 1). Breastfeeding duration was also positively associated with global GM volume (p < 0.0001). We did not find any significant negative association between breastfeeding duration and localized GM volumes. To test the robustness of this result, we divided our sample in half and repeated the VBM analysis in each of the ARMS. The only cluster that appeared on both analyses at the stipulated threshold was the one involving the left pars orbitalis and the lateral orbitofrontal cortex (left POrb/lOFC cluster).

**Figure 1.**
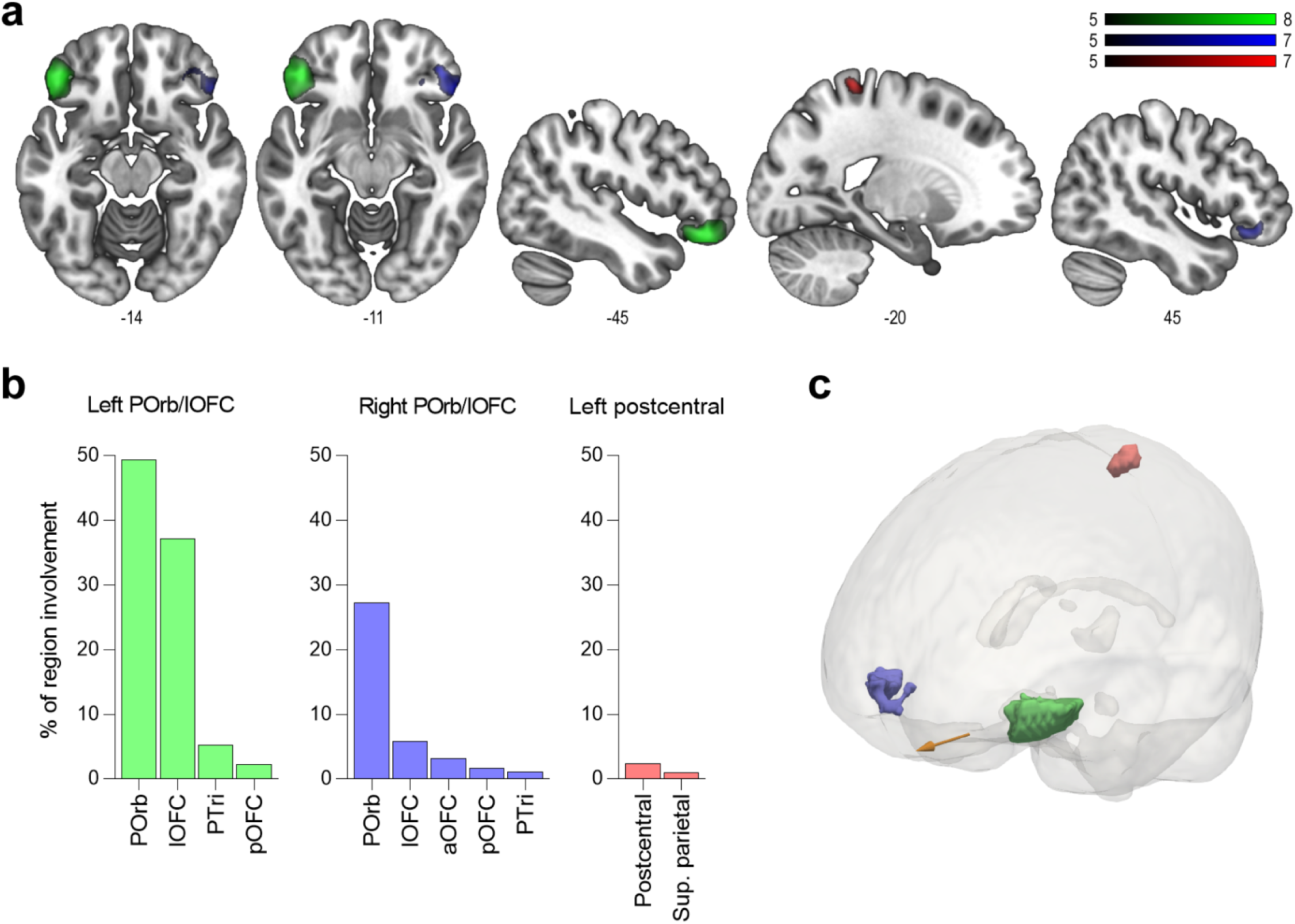
Voxel-based morphometry analysis of breastfeeding duration. **a)** The three clusters in which we found an association between breastfeeding duration and larger GM volumes are depicted. The left POrb/lOFC cluster is shown in green, the right POrb/lOFC cluster in blue, and the left postcentral/superior parietal cluster in red. The numbers below the axial and sagittal slices denote the ‘z’ and ‘x’ MNI coordinates, respectively. **b)** Histograms depicting the percentage of involvement of different brain regions—in relation to their total volume—by each of the three clusters. **c)** Three-dimensional representation of the three clusters. The orange arrow points to the anterior part of the brain. This analysis included age, sex, education level, race, ethnicity, scanner, and total intracranial volume as covariates, and we employed an FWE-corrected voxel-level threshold defined by p < 0.001 and minimum cluster size of 200 voxels. Further analyses revealed that the most robust a7ssociations were found with the left POrb/lOFC cluster. **Abbreviations:** POrb = pars orbitalis; lOFC = lateral orbitofrontal cortex; PTri = pars triangularis; pOFC = posterior orbitofrontal cortex; aOFC = anterior orbitofrontal cortex.

### Whole-brain VBM analysis controlling for potential confounds

To ensure that genetics or shared environment factors were not confounding our results, we repeated the original VBM analysis with only one randomly selected member (sibling) per family. We performed this test 10 times with a different random combination of siblings for each. The pattern of results did not change, with the left POrb/lOFC cluster being significant in all the analyses. Similarly, all the VBM analyses with additional potentially confounding factors, as well as those excluding preterm participants or employing more precise scanner covariates, yielded the same pattern of results, and the left POrb/lOFC cluster remained significant. The full list of variables that we tested may be found in Supplementary Table 1.

### Mediation analysis

We explored the potential mediatory role of the GM volume of the left POrb/lOFC cluster, which appeared to be very robust in our VBM analyses, and global GM volume, between breastfeeding duration and behavioral scores. Since it has been suggested that the pars orbitalis and the orbitofrontal cortex are associated with semantic processing,^35^ socioemotional regulation,^35,36^ and impulsivity,^36,37^ we decided to gather additional behavioral data that could provide a good estimation of these constructs. The left POrb/lOFC cluster significantly mediated the relationship between breastfeeding duration and troubling psychotic experiences (β = -0.0024, p = 0.0239) and total impulsivity (β = -0.0022, p = 0.0464). Since we found a significant result with total impulsivity, we explored all the impulsivity dimensions of the UPPS, finding an additional significant mediatory effect of the left POrb/lOFC cluster between breastfeeding duration and negative urgency (β = -0.0034, p = 0.0018). As for the analyses with global GM volume, we found this to be a significant mediator between breastfeeding duration and psychotic experiences (β = -0.0015, p = 0.0068) and total behavioral problems (β = -0.0019, p = 0.0017). Since we found a significant result with total behavioral problems, we tested the internalizing and externalizing problems scores, and global GM volume was a significant mediator in both cases (β = -0.0013, p = 0.0237 and β = -0.0020, p = 0.0009, respectively). After applying a Bonferroni correction, only the mediatory effects of the left POrb/lOFC cluster between breastfeeding duration and negative urgency, as well as those found for global GM volume and total and externalizing behavioral problems, remained significant (Figure 2). The results of all the mediation analyses may be found in Supplementary Table 3.

**Figure 2.**
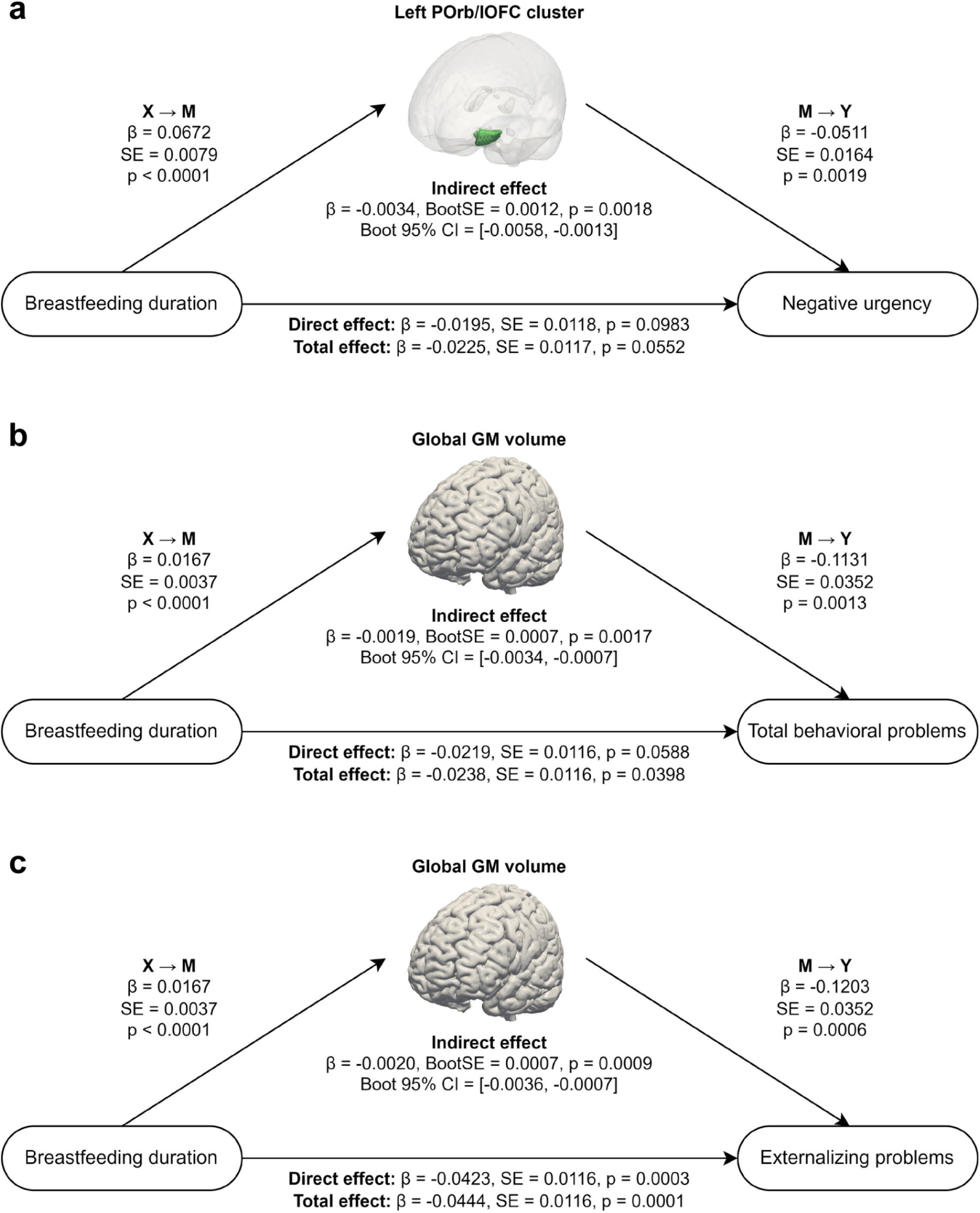
Results of the mediation analyses. **a)** We found the left POrb/lOFC cluster significantly mediated the relationship between breastfeeding duration and the negative urgency dimension of the UPPS. **b, c)** Global GM volume significantly mediated the relationship between breastfeeding duration and the total and externalizing problems scores from the Child Behavior Checklist. We report the standardized coefficients (β) and standard errors (SE). Only those mediation analyses that remained significant after Bonferroni correction are shown. The results of all the mediation analyses may be found in Supplementary Table 3.

## Discussion

In the present report we show that breastfeeding duration is mainly associated with larger bilateral volumes of the pars orbitalis and the lateral orbitofrontal cortex, as well as with larger volumes in the left postcentral gyrus and superior parietal lobule, in children aged 9 to 11 years old. Ancillary analyses demonstrated, however, that the most robust association is found in the region of the left pars orbitalis and lateral orbitofrontal. Importantly, we tested a large number of potentially confounding factors, such as birth weight, prematurity, parents’ age and education, and problems during pregnancy or birth, among many others, that did not seem to explain the relationship between breastfeeding duration and larger volumes in this region. In addition, the volume of the left pars orbitalis and lateral orbitofrontal mediates the relationship between breastfeeding duration and negative urgency impulsivity and troubling psychotic experiences. Global GM volume, for its part, also appears to mediate the association between breastfeeding duration and general behavioral problems.

To our knowledge, this is the largest study addressing the effects of breastfeeding in the brain. Most previous studies on this topic have employed very global brain measures, such as total GM volume, or have examined only a few specific brain regions, thus being more prone to obtaining less robust and less generalizable results. In this regard, our findings do not support some of these previous studies that have reported larger hippocampal volumes in association with breastfeeding.^12–14^ However, our results are partially in agreement with other studies that have utilized a whole-brain approach like the one employed here. Solis-Urra et al.^10^ reported larger volumes of the bilateral pars orbitalis associated with duration of breastfeeding in a sample of children with obesity. We have confirmed these findings in a larger sample—and largely representative of the general population—and extended them to adjacent regions. For their part, Koshiyama et al.^11^ found duration of breastfeeding to be associated with larger medial orbitofrontal volumes in children aged 10 to 13. While our main result points to a brain region close to this one, we found larger GM volumes in the lateral part of the orbitofrontal cortex, rather than the medial. Also, and similar to what we found, Ou et al.^9^ reported a small part of the left superior parietal lobule to be larger in 8-year-old children who had been breastfed in comparison with those fed with milk formula, even though this result did not prove to be statistically robust in our study. Interestingly, an experimental study with monkeys that were assigned either to a breastfeeding group or to one of two milk formula groups, concluded that breastfeeding promoted maturation of cortical GM, and this was particularly prominent in frontal regions,^17^ which is in line with our findings. While the mean age of the participants of our study was similar to that of Ou et al.,^9^ Solis-Urra et al.,^10^ and Koshiyama et al.^11^ (approximately 8, 10, and 11.6 years old, respectively), some methodological differences between studies should be noted and may serve to explain in part the variation in the results. The most obvious difference lies in the sample sizes employed; while we analyzed nearly 8,000 children, the studies of Ou et al.,^9^ Solis-Urra et al.,^10^ and Koshiyama et al.^11^ included substantially smaller numbers of participants (42, 92, and 207, respectively), which might have limited their power to identify the brain regions that we report here. Second, Ou et al.^9^ compared brain differences between breastfed children and those fed with milk formula, and Solis-Urra et al.^10^ and Koshiyama et al.^11^ employed a categorical variable for breastfeeding duration, while we assessed the effects of breastfeeding duration with a continuous measure. Lastly, other differences, such as the covariates included in the analyses, the population studied, and how data were acquired, could also partially account for these discrepancies.

Even though the exact role of the orbitofrontal cortex is still unknown,^36,38^ it is thought to be primarily involved in decision-making and in the regulation of emotions and social behavior.^36,39^ In fact, people with lesions in this region tend to be more impulsive.^36,37^ In particular, increased activity at the lateral part of the orbitofrontal cortex has been associated with less impulsive choices,^40^ while lesions in this lateral region seem to promote impulsive behavior.^41^ For its part, the pars orbitalis seems to play a role in semantic and emotional processing,^35,42^ and even one study showed that cortical thinning in this region was associated with higher impulsivity.^43^ Overall, the evidence available on the roles of these two areas appears to be largely consistent with our findings that revealed an indirect relationship between breastfeeding duration and reduced negative urgency, i.e., acting without thinking under negative emotions, through larger volumes of the left POrb/lOFC cluster. Therefore, our results suggest that breastfeeding is important for the proper development of the pars orbitalis and the lateral orbitofrontal cortex, which, in turn, appear to regulate impulsive personality traits.

Both global GM volume and the left POrb/lOFC cluster mediated the relationship between breastfeeding duration and psychotic experiences and troubling psychotic experiences, respectively. Troubling psychotic experiences, a measure of the disturbance caused by these events, are likely a better reflection of true psychotic-like experiences than just the raw number of psychotic experiences reported. Even though we deemed these two results to be non-significant after we corrected for multiple comparisons, we think that this is still a plausible finding that merits further research, for two main reasons. First, reduced volumes or surface area of both the pars orbitalis and the lateral orbitofrontal have been found to be associated with psychosis,^44–^ 47 although contradictory evidence in this respect also exists [see, for example, Lacerda et al.^48^ and Alloza et al.^49^]. Second, impulsivity—which, as noted above, seems to be indirectly affected by breastfeeding duration through the left POrb/lOFC cluster—is a common trait of patients with psychosis^50–53^ and even of unaffected relatives of these patients.^52,54^ By way of example, and bearing in mind that this is a non-clinical sample displaying a low rate of psychotic experiences, those children breastfed for more than 12 months reported having up to 36% fewer psychotic experiences and 44% lower scores for troubling experiences compared with those not breastfed or breastfed for less than 1 month, suggesting that children breastfed for longer are less prone to psychosis. Of note, some relatively dated studies tried to elucidate whether a relationship between breastfeeding and risk of developing schizophrenia existed; while two of them found breastfeeding to have a protective role,^4,5^ others did not,^55,56^ and another reported that breastfeeding just delayed the onset of the illness.^57^ Considering all of the above, the association between breastfeeding and psychosis is an intriguing line of research that needs to be addressed in a more thorough manner, ideally employing large clinical samples. While obviously breastfeeding duration will not determine whether an individual will develop a psychotic or any other mental disorder in the future, it cannot be ruled out as a potential protective factor that may be important to consider, particularly for those parents whose offspring are already at a higher risk from birth.

We found global GM volume mediated between breastfeeding duration and behavioral problems, especially externalizing behavioral problems (i.e., those specifically referring to aggressive and rule-breaking behaviors). This further supports the need to thoroughly revise the relationship between breastfeeding and mental health, as suggested in the previous paragraph. Notably, a relationship between breastfeeding duration and behavioral problems—and, in particular, the externalizing ones—had already been reported using the same behavioral test (Child Behavior Checklist) we used here.^3^ In addition, previous studies have reported beneficial effects of breastfeeding on cognition,^1^ including one study based on the ABCD dataset.^24^ According to the results of an experimental intervention, the largest cognitive differences are found in verbal measures.^58,59^ While we indeed observed a relationship between breastfeeding duration and cognition (including verbal cognition), it is somewhat striking that we did not find this relationship to be mediated by the left POrb/lOFC cluster, since it includes regions—particularly the pars orbitalis, which is part of the inferior frontal gyrus—that are recognized as being implicated in verbal functions. Nor did global GM volume mediate this relationship. Future studies may investigate what the mediating factors are between breastfeeding and cognition, and whether there are more specific cerebral mediators between breastfeeding and behavioral problems, since global GM volume is a very unspecific factor.

It is important to note that with the available data we cannot infer the mechanisms by which breastfeeding exerts its effects on the brain. Given the positive effect that breast milk seems to have on enhancing the immune system and regulating the gut microbiota,^60^ two entities that are closely interconnected,^61^ we may speculate that infants that are breastfed for longer find it easier to deal with potential challenges during the brain development period thanks to a more efficient immune system with which to fight infections and/or a proper functioning gut-brain axis. However, this remains an open question that should be further investigated.

The present study has several strengths. First, we employed a very large sample, largely representative of the United States population, thereby making our results more generalizable. Second, we used a whole-brain approach and a continuous measure of breastfeeding duration, allowing us to reliably and in an unbiased manner locate the effects of breastfeeding on the brain. Third, we controlled for several potentially confounding factors to verify that our effects were actually explained by breastfeeding duration. Conversely, we also must note some limitations. The main one is the lack of information about the exact composition and amount of breast milk taken in by the participants in their infancy. Moreover, we did not have information as to whether the participants were exclusively breastfed or were also fed with milk formula. An additional limitation is that biological mothers had to recall the number of months that they were breastfeeding, something that occurred approximately 10 years before, which is a potential source of error. However, one study showed that breastfeeding duration is quite precisely recalled even after 20 years.^62^ Finally, we could not remove the potentially confounding effects of care practices themselves (skin-to-skin contact or “kangaroo care”) and the amount of time devoted to the infant, since we did not have this information. We nevertheless considered the current degree of parental monitoring and attentiveness, which may have provided a decent estimation of the degree of attachment between parents and their infants in the past.

In conclusion, breastfeeding seems to be a major factor in the maturation of the brain, particularly for the proper development of the pars orbitalis and the lateral orbitofrontal cortex. In turn, this seems to have important consequences on the personality and mental health of the future child.

## Supporting information

STROBE

## Data Availability

All data used in the present study are available online at https://abcdstudy.org

## Acknowledgments

Data used in the preparation of this article were obtained from the Adolescent Brain Cognitive Development^SM^ (ABCD) Study (https://abcdstudy.org), held in the NIMH Data Archive (NDA). This is a multisite, longitudinal study designed to recruit more than 10,000 children age 9-10 and follow them over 10 years into early adulthood. The ABCD Study® is supported by the National Institutes of Health and additional federal partners under award numbers U01DA041048, U01DA050989, U01DA051016, U01DA041022, U01DA051018, U01DA051037, U01DA050987, U01DA041174, U01DA041106, U01DA041117, U01DA041028, U01DA041134, U01DA050988, U01DA051039, U01DA041156, U01DA041025, U01DA041120, U01DA051038, U01DA041148, U01DA041093, U01DA041089, U24DA041123, U24DA041147. A full list of supporters is available at https://abcdstudy.org/federal-partners.html. A listing of participating sites and a complete listing of the study investigators can be found at https://abcdstudy.org/consortium_members/. ABCD consortium investigators designed and implemented the study and/or provided data but did not necessarily participate in the analysis or writing of this report. This manuscript reflects the views of the authors and may not reflect the opinions or views of the NIH or ABCD consortium investigators. The ABCD data repository grows and changes over time. The ABCD data used in this report came from 10.15154/1519007. DOIs can be found at https://nda.nih.gov/study.html?id=901.

## References

1. Horta, B. L., Loret de Mola, C. & Victora, C. G. Breastfeeding and intelligence: a systematic review and meta-analysis. Acta Paediatr. 104, 14–19 (2015).

2. Victora, C. G. et al. Breastfeeding in the 21st century: epidemiology, mechanisms, and lifelong effect. Lancet 387, 475–490 (2016).

3. Oddy, W. H. et al. The long-term effects of breastfeeding on child and adolescent mental health: a pregnancy cohort study followed for 14 years. J. Pediatr. 156, 568–574 (2010).

4. McCreadie, R. G. The Nithsdale Schizophrenia Surveys. 16. Breast-feeding and schizophrenia: preliminary results and hypotheses. Br. J. Psychiatry 170, 334–337 (1997).

5. Sørensen, H. J., Mortensen, E. L., Reinisch, J. M. & Mednick, S. A. Breastfeeding and risk of schizophrenia in the Copenhagen Perinatal Cohort. Acta Psychiatr. Scand. 112, 26–29 (2005).

6. Tseng, P.-T. et al. Maternal breastfeeding and attention-deficit/hyperactivity disorder in children: a meta-analysis. Eur. Child Adolesc. Psychiatry 28, 19–30 (2019).

7. Loret de Mola, C. et al. Breastfeeding and mental health in adulthood: A birth cohort study in Brazil. J. Affect. Disord. 202, 115–119 (2016).

8. Luby, J. L., Belden, A. C., Whalen, D., Harms, M. P. & Barch, D. M. Breastfeeding and Childhood IQ: The Mediating Role of Gray Matter Volume. J. Am. Acad. Child Adolesc. Psychiatry 55, 367–375 (2016).

9. Ou, X. et al. Voxel-Based Morphometry and fMRI Revealed Differences in Brain Gray Matter in Breastfed and Milk Formula-Fed Children. AJNR Am. J. Neuroradiol. 37, 713–719 (2016).

10. Solis-Urra, P. et al. Early life factors, gray matter brain volume and academic performance in overweight/obese children: The ActiveBrains project. Neuroimage 202, 116130 (2019).

11. Koshiyama, D. et al. Association between duration of breastfeeding based on maternal reports and dorsal and ventral striatum and medial orbital gyrus volumes in early adolescence. Neuroimage 220, 117083 (2020).

12. Higgins, R. C. et al. Influence of exclusive breastfeeding on hippocampal structure, satiety responsiveness, and weight status. Matern. Child Nutr. e13333 (2022).

13. Belfort, M. B. et al. Breast Milk Feeding, Brain Development, and Neurocognitive Outcomes: A 7-Year Longitudinal Study in Infants Born at Less Than 30 Weeks’ Gestation. J. Pediatr. 177, 133–139.e1 (2016).

14. Ottolini, K. M., Andescavage, N., Kapse, K., Jacobs, M. & Limperopoulos, C. Improved brain growth and microstructural development in breast milk-fed very low birth weight premature infants. Acta Paediatr. 109, 1580–1587 (2020).

15. Blesa, M. et al. Early breast milk exposure modifies brain connectivity in preterm infants. Neuroimage 184, 431–439 (2019).

16. Isaacs, E. B. et al. Impact of breast milk on intelligence quotient, brain size, and white matter development. Pediatr. Res. 67, 357–362 (2010).

17. Liu, Z. et al. The effects of breastfeeding versus formula-feeding on cerebral cortex maturation in infant rhesus macaques. Neuroimage 184, 372–385 (2019).

18. Deoni, S. C. L. et al. Breastfeeding and early white matter development: A cross-sectional study. Neuroimage 82, 77–86 (2013).

19. Ou, X. et al. Sex-specific association between infant diet and white matter integrity in 8-y-old children. Pediatr. Res. 76, 535–543 (2014).

20. Bauer, C. E. et al. Breastfeeding Duration Is Associated with Regional, but Not Global, Differences in White Matter Tracts. Brain Sci 10, (2019).

21. Kar, P. et al. Association between breastfeeding during infancy and white matter microstructure in early childhood. Neuroimage 236, 118084 (2021).

22. Kafouri, S. et al. Breastfeeding and brain structure in adolescence. Int. J. Epidemiol. 42, 150–159 (2013).

23. Garavan, H. et al. Recruiting the ABCD sample: Design considerations and procedures. Dev. Cogn. Neurosci. 32, 16–22 (2018).

24. Lopez, D. A. et al. Breastfeeding Duration Is Associated With Domain-Specific Improvements in Cognitive Performance in 9-10-Year-Old Children. Front Public Health 9, 657422 (2021).

25. Casey, B. J. et al. The Adolescent Brain Cognitive Development (ABCD) study: Imaging acquisition across 21 sites. Dev. Cogn. Neurosci. 32, 43–54 (2018).

26. Wilke, M., Altaye, M., Holland, S. K. & CMIND Authorship Consortium. CerebroMatic: A Versatile Toolbox for Spline-Based MRI Template Creation. Front. Comput. Neurosci. 11, 5 (2017).

27. Gershon, R. C. et al. NIH toolbox for assessment of neurological and behavioral function. Neurology 80, S2–6 (2013).

28. Achenbach, T. M. & Rescorla, L. Manual for the ASEBA School-age Forms & Profiles: An Integrated System of Multi-informant Assessment. (ASEBA, 2001).

29. Karcher, N. R. et al. Assessment of the Prodromal Questionnaire-Brief Child Version for Measurement of Self-reported Psychoticlike Experiences in Childhood. JAMA Psychiatry 75, 853–861 (2018).

30. Gershon, R. C. et al. Language measures of the NIH Toolbox Cognition Battery. J. Int. Neuropsychol. Soc. 20, 642–651 (2014).

31. Goodman, R., Meltzer, H. & Bailey, V. The Strengths and Difficulties Questionnaire: a pilot study on the validity of the self-report version. Eur. Child Adolesc. Psychiatry 7, 125–130 (1998).

32. Watts, A. L., Smith, G. T., Barch, D. M. & Sher, K. J. Factor structure, measurement and structural invariance, and external validity of an abbreviated youth version of the UPPS-P Impulsive Behavior Scale. Psychol. Assess. 32, 336–347 (2020).

33. Hayes, A. F. Introduction to Mediation, Moderation, and Conditional Process Analysis. (The Guilford Press, 2022).

34. Falk, C. F. & Biesanz, J. C. Two Cross-Platform Programs for Inferences and Interval Estimation About Indirect Effects in Mediational Models. SAGE Open 6, 2158244015625445 (2016).

35. Belyk, M., Brown, S., Lim, J. & Kotz, S. A. Convergence of semantics and emotional expression within the IFG pars orbitalis. Neuroimage 156, 240–248 (2017).

36. Rudebeck, P. H. & Rich, E. L. Orbitofrontal cortex. Curr. Biol. 28, R1083–R1088 (2018).

37. Torregrossa, M. M., Quinn, J. J. & Taylor, J. R. Impulsivity, compulsivity, and habit: the role of orbitofrontal cortex revisited. Biol. Psychiatry 63, 253–255 (2008).

38. Stalnaker, T. A., Cooch, N. K. & Schoenbaum, G. What the orbitofrontal cortex does not do. Nat. Neurosci. 18, 620–627 (2015).

39. Watson, K. K. & Platt, M. L. Social signals in primate orbitofrontal cortex. Curr. Biol. 22, 2268–2273 (2012).

40. McClure, S. M., Laibson, D. I., Loewenstein, G. & Cohen, J. D. Separate neural systems value immediate and delayed monetary rewards. Science 306, 503–507 (2004).

41. Mar, A. C., Walker, A. L. J., Theobald, D. E., Eagle, D. M. & Robbins, T. W. Dissociable effects of lesions to orbitofrontal cortex subregions on impulsive choice in the rat. J. Neurosci. 31, 6398–6404 (2011).

42. Krautheim, J. T. et al. Emotion specific neural activation for the production and perception of facial expressions. Cortex 127, 17–28 (2020).

43. Merz, E. C., He, X., Noble, K. G. & Pediatric Imaging, Neurocognition, and Genetics Study. Anxiety, depression, impulsivity, and brain structure in children and adolescents. Neuroimage Clin 20, 243–251 (2018).

44. Guo, X. et al. Hippocampal and orbital inferior frontal gray matter volume abnormalities and cognitive deficit in treatment-naive, first-episode patients with schizophrenia. Schizophr. Res. 152, 339–343 (2014).

45. DeRosse, P. et al. Evidence from structural and diffusion tensor imaging for frontotemporal deficits in psychometric schizotypy. Schizophr. Bull. 41, 104–114 (2015).

46. Padmanabhan, J. L. et al. Correlations between brain structure and symptom dimensions of psychosis in schizophrenia, schizoaffective, and psychotic bipolar I disorders. Schizophr. Bull. 41, 154–162 (2015).

47. Jalbrzikowski, M. et al. Structural Brain Alterations in Youth With Psychosis and Bipolar Spectrum Symptoms. J. Am. Acad. Child Adolesc. Psychiatry 58, 1079–1091 (2019).

48. Lacerda, A. L. T. et al. Morphology of the orbitofrontal cortex in first-episode schizophrenia: relationship with negative symptomatology. Prog. Neuropsychopharmacol. Biol. Psychiatry 31, 510–516 (2007).

49. Alloza, C. et al. Psychotic-like experiences, polygenic risk scores for schizophrenia, and structural properties of the salience, default mode, and central-executive networks in healthy participants from UK Biobank. Transl. Psychiatry 10, 122 (2020).

50. Nolan, K. A., D’Angelo, D. & Hoptman, M. J. Self-report and laboratory measures of impulsivity in patients with schizophrenia or schizoaffective disorder and healthy controls. Psychiatry Res. 187, 301–303 (2011).

51. Reddy, L. F. et al. Impulsivity and risk taking in bipolar disorder and schizophrenia. Neuropsychopharmacology 39, 456–463 (2014).

52. Fortgang, R. G., Hultman, C. M., van Erp, T. G. M. & Cannon, T. D. Multidimensional assessment of impulsivity in schizophrenia, bipolar disorder, and major depressive disorder: testing for shared endophenotypes. Psychol. Med. 46, 1497–1507 (2016).

53. Nanda, P. et al. Impulsivity across the psychosis spectrum: Correlates of cortical volume, suicidal history, and social and global function. Schizophr. Res. 170, 80–86 (2016).

54. Ho, B.-C., Barry, A. B. & Koeppel, J. A. Impulsivity in unaffected adolescent biological relatives of schizophrenia patients. J. Psychiatr. Res. 97, 47–53 (2018).

55. Leask, S. J., Done, D. J., Crow, T. J., Richards, M. & Jones, P. B. No association between breast-feeding and adult psychosis in two national birth cohorts. Br. J. Psychiatry 177, 218–221 (2000).

56. Sasaki, T. et al. Type of feeding during infancy and later development of schizophrenia. Schizophr. Res. 42, 79–82 (2000).

57. Amore, M. et al. Can breast-feeding protect against schizophrenia? Case-control Study. Biol. Neonate 83, 97–101 (2003).

58. Kramer, M. S. et al. Breastfeeding and child cognitive development: new evidence from a large randomized trial. Arch. Gen. Psychiatry 65, 578–584 (2008).

59. Yang, S. et al. Breastfeeding during infancy and neurocognitive function in adolescence: 16-year follow-up of the PROBIT cluster-randomized trial. PLoS Med. 15, e1002554 (2018).

60. Gura, T. Nature’s first functional food. Science 345, 747–749 (2014).

61. Morais, L. H., Schreiber, H. L., 4th & Mazmanian, S. K. The gut microbiota-brain axis in behaviour and brain disorders. Nat. Rev. Microbiol. 19, 241–255 (2021).

62. Natland, S. T., Andersen, L. F., Nilsen, T. I. L., Forsmo, S. & Jacobsen, G. W. Maternal recall of breastfeeding duration twenty years after delivery. BMC Med. Res. Methodol. 12, 179 (2012).

