## Supplementary Table 1 for "Breastfeeding duration is associated with larger cortical gray matter volumes in children from the ABCD study"

**Supplementary Table 1.** Full list of variables of interest and covariates used in the voxel-based morphometry and mediation analyses.

| Variable name | N | ABCD document | ABCD variable name | Median (IQR) [Min – Max] or Frequency |
|---|---|---|---|---|
| Age (months) | 7,860 | abcd_mri01 | interview_age | 119 (14) [107 – 133] |
| Sex (male / female) | 7,860 | pdem02 | sex | 3,940 / 3,920 |
| Education level[a] | 7,860 | pdem02 | demo_ed_v2 | 4 (1) [1 – 7] |
| Race (white / black / american indian / asian / other or more than one race)[b] | 7,860 | pdem02 | demo_race_a_p___10 to demo_race_a_p___99 | 5,221 / 1,197 / 36 / 136 / 1,270 |
| Ethnicity (hispanic / not hispanic) | 7,860 | pdem02 | demo_ethn_v2 | 1,630 / 6,230 |
| Scanner (Siemens / GE / Philips)[c] | 7,860 | abcd_mri01 | mri_info_manufacturer | 5,210 / 1,700 / 950 |
| Total intracranial volume ($cm^3$)[d] | 7,860 | - | - | 1,477 (185) [1,006 – 2,014] |
| Weight at birth (kg) | 7,752 | dhx01 | birth_weight_lbs and birth_weight_oz | 3.23 (0.82) [0.82 – 6.41] |
| Premature birth (weeks)[e] | 7,828 | dhx01 | devhx_12_p | 0 (0) [0 – 13] |
| Age of mother at child's birth (years) | 7,807 | dhx01 | devhx_3_p | 30 (9) [13 – 60] |
| Age of father at child's birth (years) | 7,611 | dhx01 | devhx_4_p | 32 (9) [14 – 68] |
| Problems during pregnancy (number) | 7,691 | dhx01 | devhx_10a3_p to devhx_10m3_p | 0 (1) [0 – 13] |
| Birth complications (number) | 7,498 | dhx01 | devhx_14a3_p to devhx_14h3_p | 0 (1) [0 – 8] |
| Maternal tobacco use during pregnancy (yes / no) | 7,853 | dhx01 | devhx_9_tobacco | 343 / 7,510 |
| Maternal alcohol use during pregnancy (yes / no) | 7,843 | dhx01 | devhx_9_alcohol | 177 / 7,666 |
| Family income[f] | 7,199 | pdem02 | demo_comb_income_v2 | 8 (3) [1 – 10] |
| Maximum parental education level[g] | 7,851 | pdem02 | Maximum value of demo_prnt_ed_v2 and demo_prtnr_ed_v2 | 18 (4) [3 – 21] |
| Current degree of parental monitoring | 7,850 | pmq01 | Sum of parent_monitor_q1_y to parent_monitor_q5_y | 23 (3) [9 – 25] |

| Variable name | N | ABCD document | ABCD variable name | Median (IQR) [Min – Max] or Frequency |
|---|---|---|---|---|
| Current degree of parental attentiveness[h] | 7,841 | crpbi01 | Sum of crpbi_parent1_y to crpbi_parent5_y | 15 (2) [5 – 15] |
| Current body mass index (kg/m$^2$)[i] | 7,802 | abcd_ant01 | anthroheightcalc and anthroweightcalc | 17.58 (4.60) [12.04 – 36.88] |
| Degree of pubertal development | 6,242 | abcd_ssphp01 and abcd_ssphy01 | Sum of pds_p_ss_male_category and pds_y_ss_male_category or sum of pds_p_ss_female_category and pds_y_ss_female_category, depending on the sex of the participant | 4 (2) [2 – 9] |
| Preterm (yes / no)[j] | 7,828 | dhx01 | devhx_12_p | 1,082 / 6,746 |
| Scanner serial number (29 unique identifiers)[k] | 7,860 | abcd_mri01 | mri_info_deviceserialnumber | Number of participants on each scanner ranging from 12 to 775 |
| ABCD site (22 sites)[l] | 7,860 | abcd_lt01 | site_id_l | Number of participants on each ABCD site ranging from 16 to 772 |

[a] Education level of the participant: 0 = Kindergarten; 1 = 1st grade; 2 = 2nd grade; 3 = 3rd grade; 4 = 4th grade; 5 = 5th grade; 6 = 6th grade; 7 = 7th grade; 8 = 8th grade; 9 = 9th grade; 10 = 10th grade; 11 = 11th grade; 12 = 12th grade

[b] We used 4 dummy variables to represent race

[c] We used 2 dummy variables to represent scanner

[d] We obtained total intracranial volume from image processing

[e] Weeks of prematurity is represented by a continuous variable ranging from 0 to 13 (a value of 13 denotes more than 12 weeks of prematurity)

[f] Family income: 1 = Less than $5,000; 2 = $5,000-$11,999; 3 = $12,000-$15,999; 4 = $16,000-$24,999; 5 = $25,000-$34,999; 6 = $35,000-$49,999; 7 = $50,000-$74,999; 8 = $75,000-$99,999; 9 = $100,000-$199,999; 10 = $200,000 or greater

[g] Maximum parental education level: 0 = Never attended / Kindergarten only; 1 = 1st grade; 2 = 2nd grade; 3 = 3rd grade; 4 = 4th grade; 5 = 5th grade; 6 = 6th grade; 7 = 7th grade; 8 = 8th grade; 9 = 9th grade; 10 = 10th grade; 11 = 11th grade; 12 = 12th grade; 13 = High school graduate; 14 = GED or equivalent; 15 = Some college; 16 = Associate degree: Occupational; 17 = Associate degree: Academic program; 18 = Bachelor's degree; 19 = Master's degree; 20 = Professional School degree; 21 = Doctoral degree

[h] To calculate the current degree of parental attentiveness we only summed the items for the questions about the first caregiver

[i] For the analysis including current body mass index as a potential confounding factor, we excluded participants with body mass index values < 12 or > 37 (n = 58), to prevent including participants that could have mistaken height and/or weight values

[j] For this analysis, we excluded participants born preterm. We considered preterm participants those born before 37 gestational weeks (i.e., more than 3 weeks of prematurity)

[k] We used 28 dummy variables to represent the scanner serial number. We used these variables in an additional analysis replacing the original scanner variables

[l] We used 21 dummy variables to represent ABCD site. We used these variables only in mediation analyses
