## Supplementary Table 2 for "Breastfeeding duration is associated with larger cortical gray matter volumes in children from the ABCD study"

**Supplementary Table 2.** Provenance and summary measures of the behavioral data used for the mediation analyses.

| Variable name | N | ABCD document | ABCD variable name | Median (IQR) [Min – Max] |
|---|---|---|---|---|
| Global cognition | 7,711 | abcd_tbss01 | nihtbx_totalcomp_uncorrected | 87 (12) [49 – 117] |
| Total behavioral problems | 7,857 | abcd_cbcls01 | cbcl_scr_syn_totprob_r | 12 (19) [0 – 139] |
| Internalizing behavioral problems | 7,857 | abcd_cbcls01 | cbcl_scr_syn_internal_r | 3 (6) [0 – 51] |
| Externalizing behavioral problems | 7,857 | abcd_cbcls01 | cbcl_scr_syn_external_r | 2 (6) [0 – 49] |
| Psychotic experiences | 7,851 | pps01 | Sum of prodromal_1_y to prodromal_21_y | 1 (4) [0 – 21] |
| Troubling psychotic experiences | 7,851 | pps01 | Sum of prodromal_1b_y to prodromal_21b_y | 0 (3) [0 – 78] |
| Vocabulary | 7,770 | abcd_tbss01 | nihtbx_picvocab_uncorrected | 84 (10) [36 – 119] |
| Prosociality | 7,815 | abcd_psb01 (participants' questionnaire) / psb01 (parents' questionnaire) | Sum of prosocial_q1_y to prosocial_q3_y (abcd_psb01) and prosocial_q1_p to prosocial_q3_p (psb01) | 11 (3) [1 – 12] |
| Total impulsivity | 7,840 | abcd_upps01 | Sum of upps6_y to upps39_y | 41 (11) [20 – 73] |
| Lack of premeditation | 7,840 | abcd_upps01 | Sum of upps6_y, upps16_y, upps23_y, and upps28_y | 8 (3) [4 – 16] |
| Lack of perseverance | 7,847 | abcd_upps01 | Sum of upps15_y, upps19_y, upps22_y, and upps24_y | 7 (3) [4 – 16] |
| Negative urgency | 7,846 | abcd_upps01 | Sum of upps7_y, upps11_y, upps17_y, and upps20_y | 8 (3) [4 – 16] |
| Positive urgency | 7,847 | abcd_upps01 | Sum of upps35_y, upps36_y, upps37_y, and upps39_y | 8 (5) [4 – 16] |
| Sensation seeking | 7,847 | abcd_upps01 | Sum of upps12_y, upps18_y, upps21_y, and upps27_y | 10 (4) [4 – 16] |
