## Supplementary Table 3 for "Breastfeeding duration is associated with larger cortical gray matter volumes in children from the ABCD study"

**Supplementary Table 3.** Results of all the mediation analyses. We used breastfeeding duration as the independent variable (X), the GM volume of the left POrb/lOFC cluster volume or global GM volume as the mediator (M), and behavioral scores as the dependent variable (Y). We report the standardized coefficients (β) and standard errors (SE).

| Behavioral variable (Y) | Mediator (M) | X → M → Y (Indirect effect) | X → M | M → Y | X → Y (Direct effect) | X → Y (Total effect) |
|---|---|---|---|---|---|---|
| Global cognition | Left POrb/lOFC cluster volume | β = 0.0010<br>BootSE = 0.0010<br>p = 0.2745 | β = 0.0672<br>SE = 0.0080<br>p < 0.0001 | β = 0.0152<br>SE = 0.0141<br>p = 0.2818 | β = 0.1052<br>SE = 0.0101<br>p < 0.0001 | β = 0.1061<br>SE = 0.0101<br>p < 0.0001 |
| Global cognition | Global GM volume | β = 0.0002<br>BootSE = 0.0005<br>p = 0.6859 | β = 0.0165<br>SE = 0.0037<br>p < 0.0001 | β = 0.0116<br>SE = 0.0306<br>p = 0.7053 | β = 0.1059<br>SE = 0.0101<br>p < 0.0001 | β = 0.1061<br>SE = 0.0101<br>p < 0.0001 |
| Total behavioral problems | Left POrb/lOFC cluster volume | β = -0.0010<br>BootSE = 0.0011,<br>p = 0.3372 | β = 0.0676<br>SE = 0.0079<br>p < 0.0001 | β = -0.0153<br>SE = 0.0162<br>p = 0.3444 | β = -0.0229<br>SE = 0.0116<br>p = 0.0489 | β = -0.0238<br>SE = 0.0116<br>p = 0.0398 |
| Total behavioral problems* | Global GM volume | β = -0.0019<br>BootSE = 0.0007<br>p = 0.0017 | β = 0.0167<br>SE = 0.0037<br>p < 0.0001 | β = -0.1131<br>SE = 0.0352<br>p = 0.0013 | β = -0.0219<br>SE = 0.0116<br>p = 0.0588 | β = -0.0238<br>SE = 0.0116<br>p = 0.0398 |
| Internalizing behavioral problems | Global GM volume | β = -0.0013<br>BootSE = 0.0006<br>p = 0.0237 | β = 0.0167<br>SE = 0.0037<br>p < 0.0001 | β = -0.0769<br>SE = 0.0356<br>p = 0.0308 | β = 0.0170<br>SE = 0.0117<br>p = 0.1459 | β = 0.0157<br>SE = 0.0117<br>p = 0.1787 |
| Externalizing behavioral problems* | Global GM volume | β = -0.0020<br>BootSE = 0.0007,<br>p = 0.0009 | β = 0.0167<br>SE = 0.0037<br>p < 0.0001 | β = -0.1203<br>SE = 0.0352<br>p = 0.0006 | β = -0.0423<br>SE = 0.0116<br>p = 0.0003 | β = -0.0444<br>SE = 0.0116<br>p = 0.0001 |
| Psychotic experiences | Left POrb/lOFC cluster volume | β = -0.0016<br>BootSE = 0.0011<br>p = 0.1193 | β = 0.0676<br>SE = 0.0079<br>p < 0.0001 | β = -0.0243<br>SE = 0.0158<br>p = 0.1249 | β = -0.0534<br>SE = 0.0113<br>p < 0.0001 | β = -0.0549<br>SE = 0.0113<br>p < 0.0001 |

| Behavioral variable (Y) | Mediator (M) | X → M → Y (Indirect effect) | X → M | M → Y | X → Y (Direct effect) | X → Y (Total effect) |
|---|---|---|---|---|---|---|
| Psychotic experiences | Global GM volume | β = -0.0015<br>BootSE = 0.0007<br>$p = 0.0068$ | β = 0.0166<br>SE = 0.0037<br>$p < 0.0001$ | β = -0.0902<br>SE = 0.0344<br>$p = 0.0087$ | β = -0.0533<br>SE = 0.0113<br>$p < 0.0001$ | β = -0.0549<br>SE = 0.0113<br>$p < 0.0001$ |
| Troubling psychotic experiences | Left POrb/lOFC cluster volume | β = -0.0024<br>BootSE = 0.0011<br>$p = 0.0239$ | β = 0.0676<br>SE = 0.0079<br>$p < 0.0001$ | β = -0.0358<br>SE = 0.0161<br>$p = 0.0260$ | β = -0.0463<br>SE = 0.0115<br>$p < 0.0001$ | β = -0.0485<br>SE = 0.0115<br>$p < 0.0001$ |
| Troubling psychotic experiences | Global GM volume | β = -0.0010<br>BootSE = 0.0006,<br>$p = 0.0789$ | β = 0.0166<br>SE = 0.0037<br>$p < 0.0001$ | β = -0.0582<br>SE = 0.0350<br>$p = 0.0966$ | β = -0.0475<br>SE = 0.0115<br>$p < 0.0001$ | β = -0.0485<br>SE = 0.0115<br>$p < 0.0001$ |
| Vocabulary | Left POrb/lOFC cluster volume | β = 0.0008<br>BootSE = 0.0010<br>$p = 0.3902$ | β = 0.0667<br>SE = 0.0080<br>$p < 0.0001$ | β = 0.0120<br>SE = 0.0142<br>$p = 0.3976$ | β = 0.1243<br>SE = 0.0102<br>$p < 0.0001$ | β = 0.1251<br>SE = 0.0102<br>$p < 0.0001$ |
| Prosociality | Left POrb/lOFC cluster volume | β = 0.0020<br>BootSE = 0.0011<br>$p = 0.0606$ | β = 0.0673<br>SE = 0.0080<br>$p < 0.0001$ | β = 0.0300<br>SE = 0.0162<br>$p = 0.0643$ | β = 0.0144<br>SE = 0.0116<br>$p = 0.2144$ | β = 0.0162<br>SE = 0.0116<br>$p = 0.1613$ |
| Total impulsivity | Left POrb/lOFC cluster volume | β = -0.0022<br>BootSE = 0.0011<br>$p = 0.0464$ | β = 0.0675<br>SE = 0.0079<br>$p < 0.0001$ | β = -0.0320<br>SE = 0.0163<br>$p = 0.0496$ | β = -0.0231<br>SE = 0.0117<br>$p = 0.0484$ | β = -0.0250<br>SE = 0.0116<br>$p = 0.0319$ |
| Lack of premeditation | Left POrb/lOFC cluster volume | β = -0.0001<br>BootSE = 0.0011<br>$p = 0.9326$ | β = 0.0675<br>SE = 0.0079<br>$p < 0.0001$ | β = -0.0014<br>SE = 0.0164<br>$p = 0.9336$ | β = -0.0158<br>SE = 0.0118<br>$p = 0.1816$ | β = -0.0158<br>SE = 0.0118<br>$p = 0.1778$ |
| Lack of perseverance | Left POrb/lOFC cluster volume | β = -0.0013<br>BootSE = 0.0011<br>$p = 0.2426$ | β = 0.0672<br>SE = 0.0079<br>$p < 0.0001$ | β = -0.0190<br>SE = 0.0165<br>$p = 0.2498$ | β = 0.0215<br>SE = 0.0118<br>$p = 0.0695$ | β = 0.0204<br>SE = 0.0118<br>$p = 0.0845$ |
| Negative urgency* | Left POrb/lOFC cluster volume | β = -0.0034<br>BootSE = 0.0012<br>$p = 0.0018$ | β = 0.0672<br>SE = 0.0079<br>$p < 0.0001$ | β = -0.0511<br>SE = 0.0164<br>$p = 0.0019$ | β = -0.0195<br>SE = 0.0118<br>$p = 0.0983$ | β = -0.0225<br>SE = 0.0117<br>$p = 0.0552$ |

| Behavioral variable (Y) | Mediator (M) | X → M → Y (Indirect effect) | X → M | M → Y | X → Y (Direct effect) | X → Y (Total effect) |
|---|---|---|---|---|---|---|
| Positive urgency | Left POrb/lOFC cluster volume | β = -0.0018<br>BootSE = 0.0011<br>p = 0.0889 | β = 0.0672<br>SE = 0.0079<br>p < 0.0001 | β = -0.0273<br>SE = 0.0163<br>p = 0.0937 | β = -0.0642<br>SE = 0.0117<br>p < 0.0001 | β = -0.0658<br>SE = 0.0116<br>p < 0.0001 |
| Sensation seeking | Left POrb/lOFC cluster volume | β = 0.0004<br>BootSE = 0.0011<br>p = 0.7048 | β = 0.0672<br>SE = 0.0079<br>p < 0.0001 | β = 0.0061<br>SE = 0.0163<br>p = 0.7090 | β = 0.0178<br>SE = 0.0117<br>p = 0.1298 | β = 0.0181<br>SE = 0.0117<br>p = 0.1208 |

* Denotes significant indirect effects after Bonferroni correction [p < 0.0028 (0.05 / 18)]
